# The St. Gallen 2019 Guidelines understages the Axilla in Lobular Breast Cancer – a Population-Based Study

**DOI:** 10.1101/2021.06.08.21258532

**Authors:** Ulrik Narbe, Pär-Ola Bendahl, Mårten Fernö, Christian Ingvar, Looket Dihge, Lisa Rydén

**Affiliations:** Department of Clinical Sciences, Division of Oncology, Lund University, SE-223 81 Lund, Sweden; Department of Oncology, Växjö Central Hospital, SE-352 34 Växjö, Sweden; Department of Clinical Sciences, Division of Surgery, Lund University, SE-223 81 Lund, Sweden; Department of Surgery, Skåne University Hospital, SE-221 85 Lund, Sweden; Department of Plastic and Reconstructive Surgery, Skåne University Hospital, SE-205 02 Malmö, Sweden

**Keywords:** breast cancer, St. Gallen 2019 guidelines, lobular breast cancer, sentinel node metastases, axillary lymph node dissection, luminal A-like breast cancer

## Abstract

**Background:** The St. Gallen 2019 guidelines recommend omission of completion axillary lymph node dissection (cALND) in breast cancer patients with 1-2 sentinel lymph node (SLN) metastases regardless of histopathology. Concurrently, adjuvant chemotherapy is endorsed for luminal A-like disease with ≥4 axillary lymph node (ALN) metastases. We aimed to estimate the proportion of patients with invasive lobular cancer (ILC) and invasive ductal cancer of no special type (NST) and 1-2 SLN metastases for whom cALND would indicate need of adjuvant chemotherapy.

**Methods:** Patients with ILC and NST histopathology undergoing primary surgery 2014-2017 were identified in the Swedish National Quality Breast Cancer register. After exclusion of patients with incongruent or missing data, 1886 patients who fulfilled the St. Gallen 2019 criteria for cALND omission were included in the study cohort.

**Results:** Patients with ILC (n = 329) had a higher metastatic nodal burden and more often a luminal A-like subtype compared with NST patients (n = 1507). The prevalence of ≥ 4 ALN metastases was higher in ILC (31%) than in NST (15%), corresponding to an adjusted odds of 2.26 (95% CI 1.59-3.21). Luminal A-like breast cancers with ≥4 ALN metastases were overrepresented in ILC cases (52/281 (19%)) compared to NST cases (43/1299 (3%)), P<0.001.

**Conclusions:** Patients with ILC more often had a luminal A-like breast cancer with ≥4 ALN metastases compared with NST patients. Abstaining cALND in patients with luminal A-like ILC with 1-2 SLN metastases warrants future attention as it risks nodal understaging and hence undertreatment in one-fifth of these patients.

**Source of funding:** The authors declare no conflicts of interest. The Skåne County Councils Research and Developmental Foundation, the Governmental Funding of Clinical Research within the National Health Service (ALF), the Swedish Cancer Society, the Erling Persson Family Foundation, Kronoberg County Council’s Research and Development Foundation, The Cancer Foundation Kronoberg, The Swedish Research Council and The Swedish Breast Cancer Association. The funding agencies had no role in study design or interpretation of data.

**Previous communication:** Preliminary findings were published as part of Ulrik Narbe’s doctoral thesis.

**Trial registration:** ISRCTN registry: ISRCTN14341750

## Introduction

Invasive lobular cancer (ILC) is the second most common histological subtype of breast cancer after invasive ductal cancer of no special type (NST), and comprises approximately 10-15% of all invasive breast cancer ^1^. ILC has distinct clinicopathological and genomic features which distinguishes it from NST, and the responsiveness to systemic treatment differs between the subtypes ^2–5^. The majority of ILCs are classified as luminal A-like ^2, 3, 6^. Comparing patients with ILC and NST, the long-term overall prognosis seems to be the same, although a tendency towards higher incidence of late recurrences (>10 years of follow-up) has been seen in ILC ^4, 7, 8^.

ILC has a characteristic growth pattern, with single-files of tumor cells diffusely infiltrating benign tissue ^9^. Both primary tumors and axillary lymph node (ALN) metastases tend to be nonpalpable and difficult to detect by diagnostic imaging, fine needle aspiration, and core needle biopsy ^5, 10–13^. The sensitivity of axillary ultrasound for detection of ALN metastases is lower in ILC compared to that for NST, especially in patients with a high metastatic nodal burden (three or more ALN metastases) ^11^. The clinical usefulness of magnetic resonance imaging of the breast and positron emission tomography and computed tomography (PET-CT) for pretreatment breast cancer staging in patients with ILC is not defined in the literature, and none of these techniques are included in current clinical practice ^14–16^. Patients with ILC are thus often diagnosed at a higher stage than patients with NST, tend to have a larger proportion with ≥4 ALN metastases and a higher number of non-sentinel lymph node (SLN) metastases ^17–21^. Despite these differences, current surgical and oncologic treatment guidelines are similar in patients with ILC and NST.

Presence of ALN metastases is an important negative prognostic factor in breast cancer and nodal staging is one of the cornerstones in the diagnostic work-up ^22, 23^. Until recently, the standard surgical procedure for ALN staging of clinically node negative (cN0) breast cancer has been a SLN biopsy followed by completion ALN dissection (cALND) in patients with confirmed SLN metastases. Two randomized controlled trials on ALN management, The American College of Surgical Oncology Group Z0011 trial and The International Breast Cancer Study Group 23-01 trial, have shown that, omitting cALND in patients with clinically T1-2 (≤5 cm) N0 and SLN positive breast cancer, did not affect the recurrence and survival rates during the first 10 years of follow-up ^24, 25^. Furthermore, another randomized controlled trial, the AMAROS trial, where patients with clinically T1-2N0 breast cancer and ≥1 SLN metastases undergoing breast-conserving therapy or mastectomy were randomized to either cALND or axillary radiotherapy, showed no differences in recurrence or survival rates with 10 years of follow-up ^26, 27^. The findings from these trials have led to a change of practice in the axillary management irrespective of histological subtype.

In the guidelines from the St. Gallen 2019 consensus meeting, the expert panel included all histological subtypes in the extended indication for omission of cALND. They recommend that cALND can be omitted in clinically T3 (>5 cm) N0 breast cancer with 1-2 SLN metastases, undergoing either breast-conserving therapy or mastectomy, provided that adjuvant systemic treatment and regional nodal irradiation will be delivered ^6^. In addition, these guidelines recommend that adjuvant chemotherapy should be offered to patients presenting with ≥4 ALN metastases and a luminal A-like tumor. Based on the limited number of ILC patients included in the above-mentioned studies (8%, 334/4192), the proposed differences in metastatic non-SLN involvement and a high incidence of recurrences more than 10 years past primary diagnosis, the applicability of criteria for omitting cALND in patients with ILC is still unclear ^28, 29^

The aims were therefore to compare the prevalence of ≥4 ALN metastases and the distribution of number of non-SLN metastases in ILC and NST in a large population-based cohort included in the validated Swedish National Quality Breast Cancer register ^30^ and to estimate the prevalence of luminal A-like tumors with ≥4 ALN metastases in ILC and NST, in patients meeting the criteria for omission of cALND according to the St. Gallen 2019 guidelines.

## Methods

After study approval by the Ethics Committee (2019–02139), clinicopathological characteristics were retrieved from the Swedish National Quality Breast Cancer register for patients diagnosed with primary breast cancer. The register is a validated cancer registry with a coverage of 99.99% of all breast cancers diagnosed in Sweden ^30^. The study adheres to the STROBE guidelines for observational studies ^31^. The time period 2014–2017 was chosen to ensure that key variables had a high representation and that cALND was recommended according to the Swedish Treatment Guidelines for breast cancer ^32^ in patients with SLN metastases.

### Study populations

Women diagnosed between 2014–2017 with unilateral, primary breast cancer classified as pure ILC, pure NST, or mixed ILC/NST, who underwent breast and axillary surgery as primary treatment, were identified in the Swedish National Quality Breast Cancer register (*n* = 20 139). Patients who underwent SLN biopsy and cALND, with breast cancer eligible for omission of cALND according to the St. Gallen 2019 criteria (clinically T1-3 N0 with 1-2 SLN metastases of which at least 1 macrometastasis) were included in the main study cohort (St. Gallen 2019 cohort, *n* = 1886). In addition, a Z0011 cohort was defined as patients who underwent breast-conserving therapy and SLN biopsy, in line with the Z0011 criteria for omission of cALND (clinically T1-2 N0, 1-2 SLN metastases of which at least 1 macrometastasis), *n* = 975. Patients with node-negative disease and those with data from only SLN biopsy or ALND were excluded. The flow-chart of the study is depicted in Figure 1.

**Figure 1.**
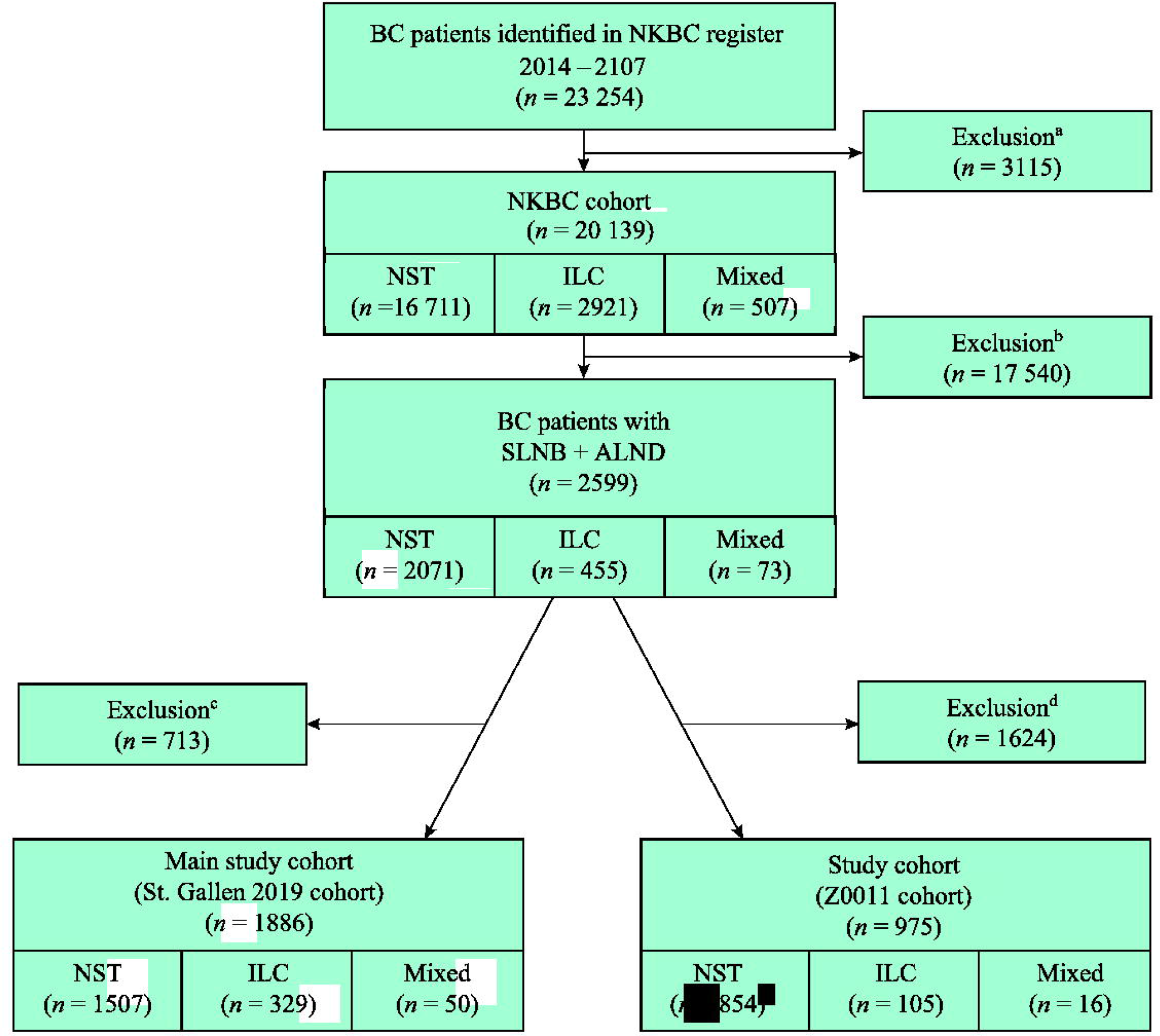
Flow-chart of the study. *Footnotes:* ^a^Nodal data incongruent or missing (*n* = 1 237), tumor size missing (*n* = 235), no invasive tumor (*n* = 33), histological subtype missing (*n* = 54), histological subtype other than ILC, NST or mixed ILC/NST (*n* = 1 865). Note that some patients were excluded for more than one reason ^b^SLNB only or ALND only ^c^T-stage 4 (T4), ≥3 SLNMs, SLN micrometastasis only ^d^T-stage 3 (T3), Mastectomy *Abbreviations:* BC: Breast cancer; NKBC: Swedish National Quality Breast Cancer Register; NST: Invasive ductal cancer of no special type; ILC: Invasive lobular cancer; SLN: Sentinel lymph node; SLNB: Sentinel lymph node biopsy; ALND: Axillary lymph node dissection

### Pathological assessment and surrogate molecular subtypes

Pathological assessments of the primary tumor, SLNs and ALNs were performed in accordance with the Swedish Quality Document for Pathology ^33^. According to the guidelines valid at the study period, ILC was identified by specified morphological criteria and in case of a histologically uncertain specimen showing lobular growth pattern a complementary immunohistochemical staining of E-cadherin was performed. ALNs were identically prepared irrespective of histological subtype, whereas SLNs were subjected to complimentary cytokeratin staining in patients with ILC and benign hematoxylin-eosin assessment. ER and progesterone receptor (PR) positivity were defined as ≥10% of immunohistochemically stained nuclei, and in accordance with Prat et al., PR ≥20% was considered high ^34^. HER2 positivity was defined as the HER2 *in situ* hybridization test positive, and if this test was missing by immunohistochemical 3+ scoring. The Ki67 percentage was categorized into 3 groups: low, intermediate and high, based on local laboratory percentile based cut-offs, and Nottingham histological grade (NHG) was evaluated according to Elston and Ellis ^35^.

In line with international guidelines, a lymph node micrometastasis was defined as a tumor deposit >0.2 mm but ≤2 mm consisting of ≥200 tumor cells, and a macrometastasis as a deposit >2 mm. Deposits ≤0.2 mm and/or consisting of <200 tumor cells were defined as isolated tumor cells. Patients with SLN isolated tumor cells only were classified as N0 ^33, 36^. Patients with 1-2 SLN metastases with SLN micrometastases only (*n* = 216, 10%) were not included in the study cohorts (Figure 1) according to current clinical guidelines for axillary surgery, where cALND is not recommended for this group of patients ^32, 37–40^.

Based on a modification of the St. Gallen 2019 guidelines ^6^ and the classification proposed by Maisonneuve et al. ^41^ (including ER, PR, HER2, Ki67 and NHG), the surrogate molecular subtypes luminal A-like; luminal B-like; HER2 positive; and triple-negative were defined. The definitions are described in Supplementary material 1.

### Statistical Analysis

The primary endpoint was prevalence of ≥4 ALN metastases in patients with ILC and NST, respectively. A secondary endpoint was to estimate the proportion of patients with luminal A-like tumors with ≥4 ALN metastases in ILC and NST, in patients meeting the criteria for omission of cALND according to the St. Gallen 2019 guidelines.

Evidence for differences in categorical variables, including patient and tumor characteristics, between the histological subtypes (ILC vs. NST) was evaluated using Pearsons’s chi-squared test, Fisher’s exact test if one or more of the expected counts in the contingency table <5, or Pearsons’s chi-squared test for trend (ordinal variables with >2 categories). Variables measured on a continuous scale were evaluated using the Mann-Whitney U-test. Uni- and multivariable analyses were performed using logistic regression.

*P*-values, which have not been adjusted for multiple testing, should be interpreted as level of evidence, on a continuous scale from zero to one, against the null hypothesis without reference to a cut-off for significance.

IBM SPSS Statistics (version 25.0, IBM Inc., Armonk, NY, USA) and STATA version 16 (StataCorp, College Station, TX, USA) were used for statistical calculations.

## Results

### Demographics and non-sentinel node metastases by histological subtype in the St. Gallen 2019 cohort

In the St. Gallen 2019 cohort, comparing 329 patients with pure ILC and 1 507 with pure NST with 1-2 metastatic SLNs, several differences in clinicopathological characteristics were identified (Table 1). Patients with ILC were older, had a lower detection rate with screening mammography and more often had mastectomy. Their tumors were larger, more often multifocal and the metastatic burden in ALNs higher. Additionally, the proportion of luminal A-like tumors was higher, while other surrogate subtypes were less frequent in ILC compared to NST. The characteristics of the mixed ILC/NST group (*n* = 50) are reported separately and the biomarker profile was closer to pure ILC than to pure NST.

**Table 1.** Clinicopathological characteristics in patients eligible for omission of completion ALND according to the St. Gallen 2019 International Consensus Guidelines

| St. Gallen 2019 eligible breast cancer<br>BCT or mastectomy, T1-3, cN0,<br>1-2 SLN metastases of which $\geq 1$ macrometastases | NST<br>(*n* = 1507)<br>*n* (%) | ILC<br>(*n* = 329)<br>*n* (%) | *P*-value<br>ILC vs. NST | Mixed ILC/NST<br>(*n* = 50)<br>*n* (%) |
| --- | --- | --- | --- | --- |
| Age at surgery: years, median (IQR) | 62 (52-71) | 65 (54-72) | 0.011<sup>b</sup> | 60 (48-70) |
| Detected by screening mammography |  |  |  |  |
| No | 799 (53) | 180 (55) | 0.549<sup>a</sup> | 25 (50) |
| Yes | 707 (47) | 148 (45) |  | 25 (50) |
| Missing | 1 | 1 |  | 0 |
| Type of breast surgery |  |  |  |  |
| BCT | 861 (57) | 113 (34) | <0.001<sup>a</sup> | 17 (34) |
| Mastectomy | 646 (43) | 226 (66) |  | 33 (66) |
| Multifocality |  |  |  |  |
| No | 1142 (76) | 233 (69) | 0.012<sup>a</sup> | 28 (57) |
| Yes | 362 (24) | 99 (31) |  | 21 (43) |
| Missing | 3 | 7 |  | 1 |
| T-stage |  |  |  |  |
| T1 ($\leq 20$ mm) | 832 (55) | 85 (26) | <0.001<sup>a</sup> | 19 (38) |
| T2 (>20 to 50 mm) | 627 (42) | 175 (53) |  | 25 (50) |
| T3 (>50 mm) | 48 (3) | 69 (21) |  | 6 (12) |
| No. of SLNs excised: median (range) | 2 (1-9) | 2 (1-8) | 0.025<sup>b</sup> | 2 (1-9) |
| No. of SLNMs |  |  |  |  |
| 1 | 1148 (76) | 243 (74) | 0.374<sup>a</sup> | 38 (76) |
| 2 | 359 (24) | 86 (26) |  | 12 (24) |
| No. of LNs excised: median (IQR) | 13 (10-17) | 12 (10-16) | 0.362<sup>b</sup> | 12 (1-16) |
| No. of non-SLNMs |  |  |  |  |
| No (0) | 962 (64) | 164 (50) | <0.001<sup>a</sup> | 31 (61) |
| Yes ($\geq 1$) | 545 (36) | 165 (50) | | 19 (39) |
| 1 | 250 (17) | 42 (13) | <0.001<sup>a</sup> | 8 (16) |
| 2 | 107 (7) | 29 (9) |  | 6 (12) |
| $\geq 3$ | 188 (12) | 94 (29) | | 5 (10) |
| 1 to 2 | 357 (24) | 71 (22) | <0.001<sup>a</sup> | 14 (28) |
| $\geq 3$ | 188 (12) | 94 (96) | | 5 (10) |
| Median (IQR) if >0 | 2 (1-3) | 3 (1-7) | <0.001<sup>b</sup> | 2 (1-3) |
| Nodal stage: 3 groups |  |  |  |  |
| N1 (1 to 3 ALNMs) | 1283 (85) | 227 (69) | <0.001<sup>a</sup> | 43 (86) |
| N2 (4 to 9 ALNMs) | 181 (12) | 69 (21) |  | 6 (12) |
| N3 ($\geq 10$ ALNMs) | 43 (3) | 33 (10) | | 1 (2) |
| Nodal stage: 2 groups |  |  |  |  |
| N1 (1 to 3 ALNMs) | 1283 (85) | 227 (69) | <0.001<sup>a</sup> | 43 (86) |
| N$\geq 2$ ($\geq 4$ ALNMs) | 224 (15) | 102 (31) | | 7 (14) |
| Molecular subtype (all patients)<sup>c</sup> |  |  |  |  |
| Luminal A-like | 484 (37) | 176 (63) | <0.001<sup>a</sup> | 26 (62) |
| Luminal B-like | 501 (39) | 88 (31) |  | 14 (33) |
| HER2 positive | 210 (16) | 13 (5) |  | 2 (5) |
| Triple-negative | 104 (8) | 4 (1) |  | 0 (0) |
| Missing | 208 | 48 |  | 8 |
| Molecular Subtype (patients with $\geq 4$ ALNMs) | | | | |
| Luminal A-like | 43 (23) | 52 (60) | <0.001<sup>a</sup> | 2 (33) |
| Luminal B-like | 91 (48) | 25 (29) |  | 4 (67) |
| HER2 positive | 42 (22) | 7 (8) |  | 0 (0) |
| Triple-negative | 15 (8) | 3 (3) |  | 0 (0) |
| Missing | 33 | 15 |  | 1 |
| Luminal A-like and $\geq 4$ ALNMs | | | | |
| No | 1256 (97) | 229 (81) |  | 40 (95) |
| Yes | 43 (3) | 52 (19) | $<0.001^a$ | 2 (5) |
| Missing | 208 | 48 |  | 8 |
| Luminal A-like |  |  |  |  |
| $<4$ ALNMs | 441 (91) | 124 (70) | $<0.001^a$ | 24 (92) |
| $\geq 4$ ALNMs | 43 (9) | 52 (30) | | 2 (8) |
| Luminal B-like |  |  |  |  |
| $<4$ ALNMs | 410 (82) | 63 (72) | $0.026^a$ | 10 (71) |
| $\geq 4$ ALNMs | 91 (18) | 25 (28) | | 4 (29) |
| HER2 positive |  |  |  |  |
| $<4$ ALNMs | 168(80) | 6 (46) | $0.004^a$ | 2 (100) |
| $\geq 4$ ALNMs | 42 (20) | 7 (54) | | 0 (0) |
| Triple-negative |  |  |  |  |
| $<4$ ALNMs | 89 (86) | 1 (25) | $0.001^a$ | 0 (0) |
| $\geq 4$ ALNMs | 15 (14) | 3 (75) | | 0 (0) |
| Subtype missing |  |  |  |  |
| $<4$ ALNMs | 175 (84) | 33 (69) | $0.014^a$ | 7 (88) |
| $\geq 4$ ALNMs | 33 (16) | 15 (31) | | 1 (12) |

<sup>a</sup>P-value from Pearson's chi-square test (linear by linear association test if more than 2 ordered categories)
<sup>b</sup>P-value from Mann-Whitney U-test
<sup>c</sup>Based on a modification of the St. Gallen 2019 guidelines and the classification proposed by Maisonneuve et al. the tumors were defined as: luminal A-like if (1) ER+, HER2-, NHG 1 or (2) ER+, HER2-, NHG 2, Ki67 low or (3) ER+, HER2-, NHG 2, Ki67 intermediate and PR $\geq 20\%$ ; luminal B-like if (1) ER+, HER2-, NHG 3 or (2) ER+, HER2-, NHG 2, Ki67 high or (3) ER+, HER2-, NHG 2, Ki67 intermediate and PR $<20\%$ ; HER2 positive (all HER2+ independent of ER, NHG, Ki67 and PR status); triple-negative (ER-, PR- and HER2-)
*Abbreviations:* ALNMs = axillary lymph node metastases, ALND = axillary lymph node dissection, LN = lymph node, SLN = sentinel lymph node, SLNMs = sentinel lymph node metastases, BCT = breast-conservation therapy, NST = invasive ductal cancer of no special type, ILC = invasive lobular cancer, IQR = interquartile range, cN0 = clinically node negative, ER = estrogen receptor, PR = progesterone receptor, HER2 = human epidermal growth factor receptor 2, NHG = Nottingham histological grade, Ki67 = proliferation marker

The number of excised SLNs and ALNs did not differ by histological subtype (Table 1). However, one or more non-SLN metastases in the axillary specimen was more common in ILC (165/329 (50%)) than in NST (545/1507 (36%)), *P* <0.001; odds ratio (OR) 1.78, 95% confidence interval (CI) 1.40-2.26) and the same pattern was observed in patients with ≥4 ALN metastases (ILC: 102/329 (31%) and NST: 224/1507 (15%)), *P* <0.001; OR 2.57, 95% CI 1.96-3.38 (Table 1) (Figure 2a,3a). In patients with one or more non-SLN metastases, the total number of non-SLN metastases was also higher in ILC compared to NST (median (interquartile range (IQR)): 3 (1-7) vs. 2 (1-3)) (Table 1) (Figure 2b).

**Figure 2.**
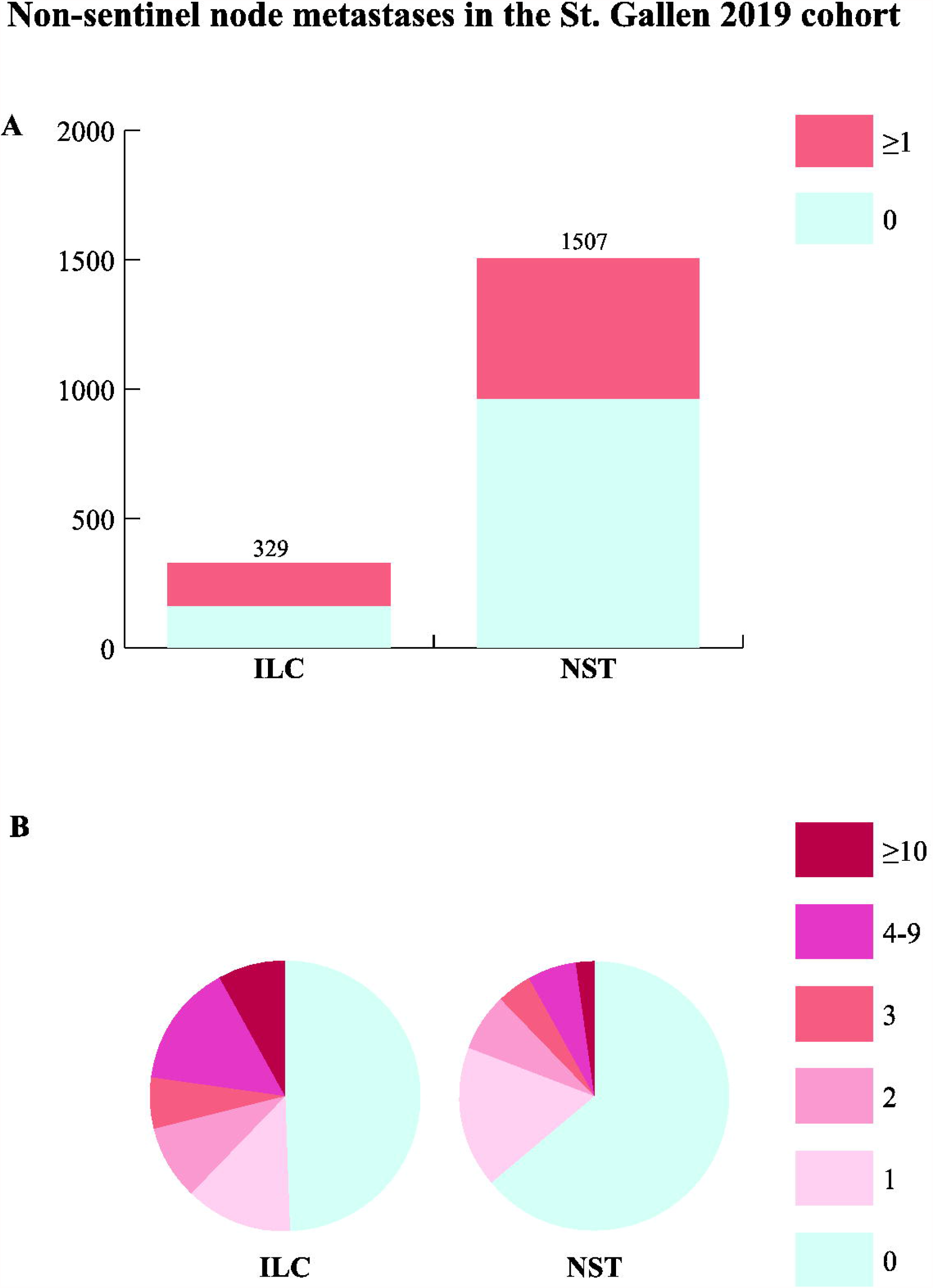
Non-sentinel node metastases in patients with invasive lobular cancer (ILC) and invasive ductal carcinoma of no special type (NST), 1-2 sentinel lymph node metastases, fulfilling the St. Gallen 2019 criteria for omission of completion axillary lymph node dissection. **A** Histological subtype in relation to the risk of non-sentinel node metastases stratified in 0 and ≥1 metastases. **B** Pie chart illustrating the proportion of 0 - ≥10 non-sentinel node metastases within ILC and NST.

### Four or more nodal metastases by histological and surrogate subtype in the St. Gallen 2019 cohort

Patients with luminal A-like subtype and four or more ALN metastases were overrepresented in ILC (*n* = 52/281 (19%)) compared to the NST cases (*n* = 43/1299 (3%)), *P* <0.001; OR 6.63 95% CI 4.32-10.2) (Table 1) (Figure 3b). Moreover, the relative frequencies of ≥4 ALN metastases within all the different surrogate molecular subtypes were also higher in ILC compared to NST cases in the St. Gallen cohort (Table 1, Figure 3C).

**Figure 3.**
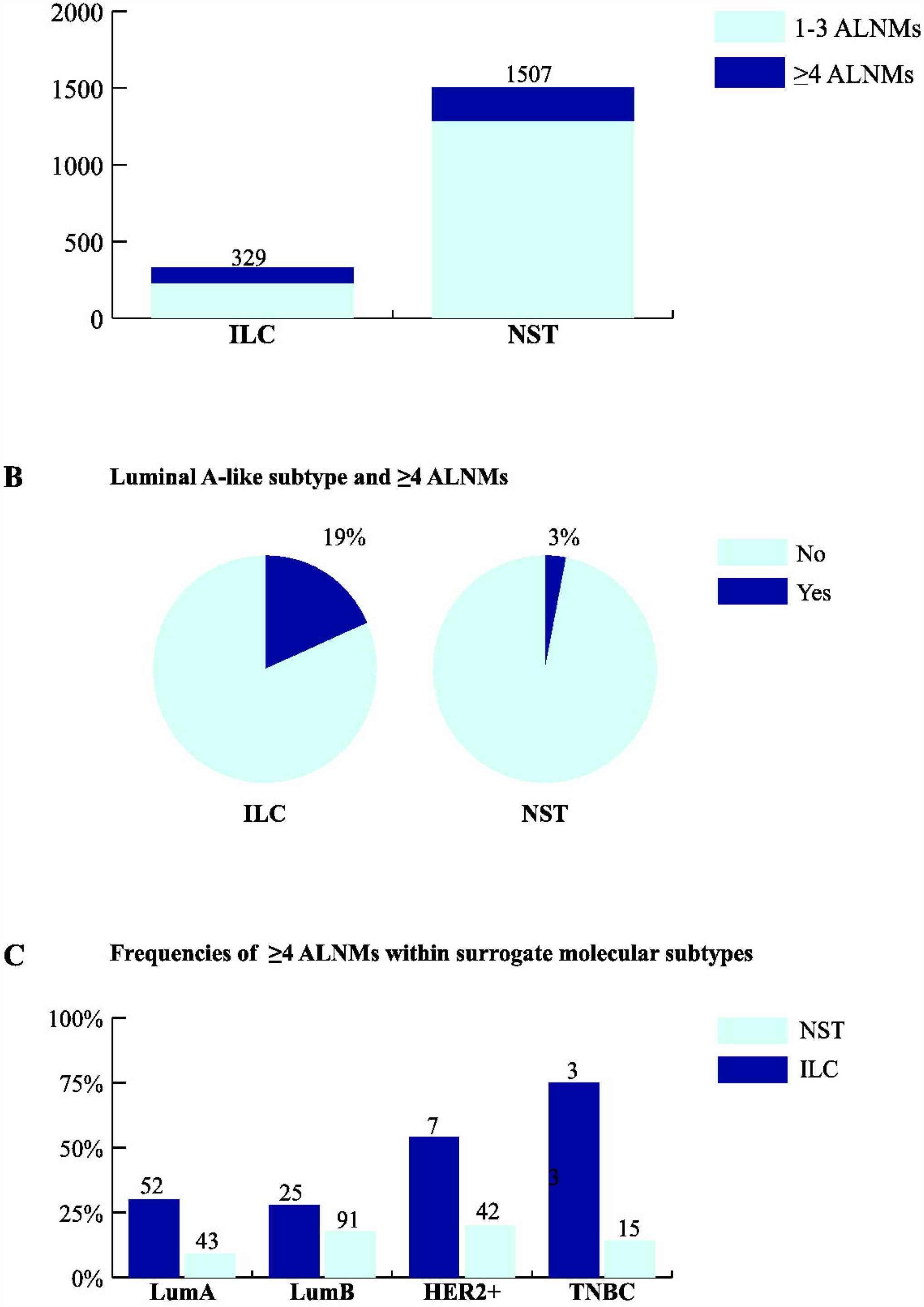
Four or more axillary lymph node metastases (ALNMs) in patients with invasive lobular carcinoma (ILC) and invasive ductal carcinoma of no special type (NST), 1-2 sentinel lymph node metastases, fulfilling the St. Gallen 2019 criteria for omission of completion axillary lymph node dissection. **A** Histological subtype in relation to ALNMs stratified into nodal stages 1-3 and ≥4 ALNMs. **B** Pie chart illustrating the proportion of patients with luminal A-like subtype and ≥4 ALNMs within ILC and NST. **C** The relative frequencies of ≥4 ALNMs within the different surrogate molecular subtypes in ILC and NST cases. *Abbreviations:* LumA: Luminal A□like; LumB: Luminal B□like; HER2+: HER2□positive; TNBC: Triple□negative

Additionally, the adjusted odds of ≥4 ALN metastases in patients with luminal A-like subtype were higher in ILC cases compared to NST cases (OR 2.92 (95% CI 1.73-4.94).

### Multivariable analyses of axillary nodal burden in the St. Gallen 2019 cohort

The odds of one or more non-SLN metastases was higher for ILC than for NST also after adjustment for other relevant predictors including age, detection by screening mammography, T-stage, multifocality, number of SLNs with macrometastases and surrogate molecular subtypes (OR 1.55, 95% CI 1.15-2.08, *P* = 0.004) (Table 2).

**Table 2.** Multivariable analysis including patients eligible for omission of completion ALND according to the St. Gallen 2019 International Consensus Guidelines^a^ (*n* = 1610)

| Variables | Non-SLN metastases yes/no | | ALN metastases $\geq 4$ vs. $< 4$ | |
| --- | --- | --- | --- | --- |
|  | OR (95% CI) | *P*-value<sup>b</sup> | OR (95% CI) | *P* value |
| Histological subtype | | 0.010<sup>c</sup> | | $< 0.001^c$ |
| ILC vs. NST | 1.55 (1.15-2.08) | 0.004 | 2.26 (1.59-3.21) | $< 0.001$ |
| Mixed ILC/NST vs. NST | 0.86 (0.44-1.68) | 0.659 | 0.85 (0.34-2.15) | 0.739 |
| Age (per year) | 1.00 (1.00-1.01) | 0.271 | 1.01 (1.00-1.02) | 0.123 |
| Detection by screening mammography |  |  |  |  |
| Yes vs. No | 0.88 (0.71-1.10) | 0.262 | 0.77 (0.57-1.03) | 0.078 |
| T-stage<sup>d</sup> | | $< 0.001^c$ | | $< 0.001^c$ |
| T2 vs. T1 | 1.40 (1.11-1.75) | 0.004 | 1.80 (1.31-2.46) | $< 0.001$ |
| T3 vs. T1 | 3.14 (1.96-5.04) | $< 0.001$ | 3.30 (1.96-5.54) | $< 0.001$ |
| Multifocality |  |  |  |  |
| Yes vs. No | 1.46 (1.15-1.86) | 0.002 | 1.37 (1.01-1.87) | 0.044 |
| SLN metastases | | $< 0.001^c$ | | $< 0.001^c$ |
| 2 (1 macro) vs. 1 (macro) | 0.76 (0.47-1.22) | 0.251 | 1.11 (0.60-2.04) | 0.744 |
| 2 (both macro) vs. 1 (macro) | 1.84 (1.41-2.39) | $< 0.001$ | 3.39 (2.50-4.50) | $< 0.001$ |
| Surrogate molecular subtypes<sup>e</sup> |  | 0.070<sup>c</sup> |  | 0.010<sup>c</sup> |
| Luminal B-like vs. Luminal A-like | 1.31 (1.03-1.66) | 0.030 | 1.51 (1.09-2.09) | 0.013 |
| HER2 positive vs. Luminal A-like | 1.43 (1.32-1.99) | 0.034 | 1.97 (1.28-3.04) | 0.002 |
| Triple-negative vs. Luminal A-like | 1.05 (0.66-1.65) | 0.844 | 1.17 (0.64-2.17) | 0.608 |

<sup>a</sup>St. Gallen 2019 International Consensus Guidelines for omission of completion ALND: T1-3, cN0, 1-2 SLN metastases of which $\geq 1$ macrometastasis ( $> 2$ mm tumor deposit), BCT or mastectomy
<sup>b</sup>*P*-value from logistic regression
<sup>c</sup>Overall test of the factor
<sup>d</sup>T-stage: T1 = $\leq 20$ mm, T2 = $> 20$ -50mm, T3 = $> 50$ mm
<sup>e</sup>Based on a modification of the St. Gallen 2019 guidelines and the classification proposed by Maisonneuve et al. the tumors were defined as: luminal A-like if (1) ER+, HER2-, NHG 1 or (2) ER+, HER2-, NHG 2, Ki67 low or (3) ER+, HER2-, NHG 2, Ki67 intermediate and PR $\geq 20\%$ ; luminal B-like if (1) ER+, HER2-, NHG 3 or (2) ER+, HER2-, NHG 2, Ki67 high or (3) ER+, HER2-, NHG 2, Ki67 intermediate and PR $< 20\%$ ; HER2 positive (all HER2+ independent of ER, NHG, Ki67 and PR status); triple-negative (ER-, PR- and HER2-)
Abbreviations: CI = confidence interval, ALN = axillary lymph node, ALND = axillary lymph node dissection, BCT = breast conservation therapy, cN0 = clinically node negative, ER = estrogen receptor, HER2 = human epidermal growth factor receptor 2, SLN = sentinel lymph node, Ki67 = proliferation marker

Furthermore, the odds of ≥4 ALN metastases was higher in ILC than in NST after adjustment for the same variables as above (OR 2.26, 95% CI 1.59-3.21, *P* <0.001) (Table 2, Figure 4). Tumor stage, multifocality, surrogate subtypes and number of SLNs with macrometastases were confirmed as predictors of ≥4 ALN metastases along with ILC (Table 2) (Figure 4). The luminal B-like and HER2 positive surrogate subtypes vs. luminal A-like were predictors of axillary nodal burden, whereas the low prevalent mixed ILC/NST histological subtype added no information on the risk of ≥4 ALN metastases (Table 2 and Figure 4).

**Figure 4.**
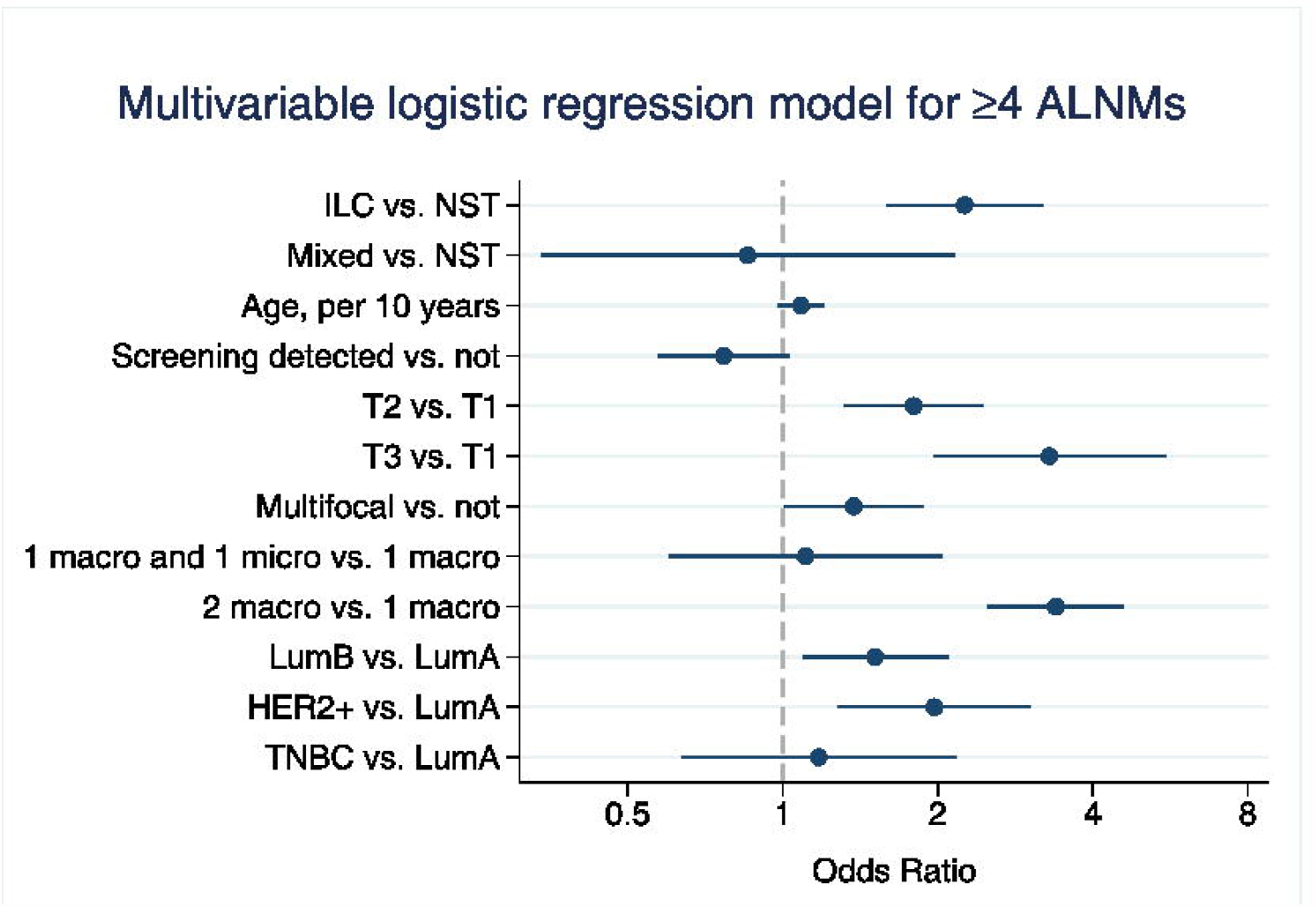
Forest plot visualization of the multivariable logistic regression model for ≥4 axillary lymph node metastases (ALNMs) in patients fulfilling the St. Gallen 2019 criteria for omission of completion axillary lymph node dissection. *Abbreviations:* ILC: Invasive lobular cancer; NST: Invasive ductal cancer of no special type; LumA: Luminal A-like; LumB: Luminal B-like; HER2+: HER2 positive; TNBC: Triple-negative breast cancer

### Demographics and nodal metastatic burden in the Z0011 cohort

Clinicopathological data from the Z0011 cohort (Supplementary material 2) and the entire cohort retrieved from the register (Supplementary material 3) showed essentially the same differences between pure ILC and pure NST cases as in the St. Gallen 2019 cohort.

In the Z0011 cohort (ILC; *n* = 105, vs. NST; *n* = 854), similar distributions of patients with one or more non-SLN metastases stratified by histological subtype was noted (ILC 42% and NST 31%) *P* = 0.02; OR 1.60, 95% CI 1.06-2.43), and the proportion of patients with ≥4 ALN metastases was also higher in ILC (ILC 26% and NST 11%), P <0.001; OR 2.90, 95% CI 1.78-4.73). Furthermore, the total number of non-SLN metastases in those with one or more non-SLNMs was higher in ILC compared with NST (Supplementary material 2).

In line with the results in the St. Gallen 2019 cohort, patients in the Z0011 cohort with luminal A-like subtype and ≥4 ALN metastases were also overrepresented in ILC (*n* = 13/83 (16%)) compared to the NST cases (*n* = 21/738 (3%)), *P* <0.001; OR 6.34, 95% CI 3.04-13.2) (Supplementary material 2).

In multivariable analyses predicting non-SLN metastases, ILC remained an independent predictor after adjustment (OR 1.81, 95% CI 1.11-2.98, *P* = 0.018) (Supplementary material 4). Finally, the adjusted odds of ≥4 ALN metastases was also higher in ILC than in NST (OR 2.94, 95% CI 1.56-5.54, *P* = 0.001) (Supplementary material 4).

## Discussion

This population based Swedish registry study shows that when applying the St. Gallen 2019 guidelines for omitting cALND, the presence of non-SLN metastases and the proportion of ≥4 ALN metastases were higher in ILC than in NST. The present study highlights that the St. Gallen 2019 criteria for omission of cALND is associated with an understaging of ALN status, and a subsequent risk for adjuvant systemic undertreatment in the clinic, especially in ILCs with luminal A-like subtype. Most patients with NST had benign non-SLN after cALND, while in ILC, half of the patients had non-SLN involvement in the axillary specimen. Similar results were seen for the narrower but more generally accepted Z0011 criteria for abstaining cALND. Importantly, both in the St. Gallen 2019 and Z0011 cohorts, ILC was an independent predictor of non-SLN metastases and ≥4 ALN metastases after adjustment for validated predictors of non-sentinel node metastases. Previous studies have included a limited number of ILC and not specifically addressed residual axillary nodal involvement after abstaining cALND when applying the St. Gallen 2019 guidelines.

Breast cancer classified as luminal A-like with ≥4 ALN metastases was more frequent in patients with ILC compared to NST (St. Gallen 2019 cohort: *n* = 52 (19%) vs. *n* = 43 (3%), Z0011 cohort: *n* = 13, 16% vs. *n* = 21, 3%, respectively). In patients with a luminal A-like subtype, treatment recommendation on adjuvant chemotherapy depends on ALN staging according to the St. Gallen 2019 consensus guidelines even in the era of genomic testing. Although patients with the luminal B-like and HER2 positive subtypes had a higher risk of ≥4 ALN metastases than patients with the luminal A-like subtype, the staging information from cALND is of less importance for these patients as they would be recommended adjuvant chemotherapy irrespective of nodal status. Our results suggest that when applying the St. Gallen 2019 criteria for omitting cALND, approximately 1 out of 5 ILC and 1 out of 30 NST patients with 1-2 confirmed SLN macrometastases will not be offered adjuvant chemotherapy as a result of understaging of the axilla, and essentially the same results were seen in the Z0011 cohort. Moreover, according to Swedish Treatment Guidelines ^32^, all patients with ≥4 ALN metastases would have diagnostic imaging recommended for the evaluation of metastatic disease before the start of adjuvant therapy.

Previous breast cancer studies exploring the impact of nodal staging data on adjuvant treatment decision, have not specified the histological subtypes and are based on smaller cohorts than ours ^42, 43^. Aigner et al. included 132 patients and, in line with our data, found that 17% of the patients with Z0011 eligible breast cancer would have been offered a more extensive adjuvant treatment based on the information retrieved by cALND, while Stenmark et al. included 238 patients with clinically N0 breast cancers with ≥1 SLN metastases (≥1 macrometastasis) and found that in 18% of those with luminal A-like tumors four or more ALN metastases were detected after cALND.

In the present study, including a population-based unselected cohort, patients eligible for omission of cALND according to St. Gallen 2019 and Z0011, displayed a prognostically more unfavorable ALN status, with a more extensive nodal metastatic burden, compared to those originally included in the Z0011, The International Breast Cancer Study Group 23-01 and AMAROS trials. Based on current clinical guidelines the study inclusion was restricted to those patients with 1-2 SLN metastases of which at least 1 macrometastasis, and patients with isolated tumor cells only were classified as N0, and excluded ^36^. In the seminal trials, a majority of the included patients had micrometastasis only in their SLNs and isolated tumor cells were classified as micrometastasis ^44^, and ≥90% of the patients received adjuvant systemic treatment. These conditions could have affected the observed outcome data and with a follow-up time restricted to 10 years, there is still a risk for late recurrences, especially for ILC ^4, 7, 8^. Additionally, a minority of the included patients had ILC (~8%) and in none of the trials, data from subgroup analyses in ILC and NST was reported.

Studies investigating ALN status in relation to histological subtype, have shown a higher axillary metastatic burden in ILC compared to NST, but the total number of patients with ILC, strictly eligible for omission of cALND according to the Z0011 criteria and with both SLN and non-SLN data available, is low (*n* = 2–24); and complete data from those meeting the St. Gallen 2019 guidelines are for the most part either missing or not reported ^17, 19, 45–47^. While numerous prediction models have been presented to support clinical decision on cALND, only few of the models include histological or molecular subtypes ^48^.

This is one of the largest studies from a validated nationwide quality register exploring ALN status in ILC vs. NST in patients meeting the criteria for omission of cALND according to St. Gallen 2019 and Z0011. All tumor deposits were assessed according to current classification for isolated tumor cells, micro- and macrometastasis and prospectively performed ^36^. A weakness in this study was that no data was available on gross extracapsular extension or vascular invasion. Hence, these risk factors for ALN metastases could not be included in the multivariable analysis. Studies including registry data may be considered less reliable, given the risks of incomplete and misclassified data. In the present study, however, the original dataset was complete for histological subtype in all except 54 cases and data on nodal status was found to be incongruent or missing in 5% (1144/23 254) of the patients.

Mamtani et al. compared patients with ILC (*n* = 104) and NST (*n* = 709) treated according to Z0011 criteria, where cALND was omitted in those with 1–2 SLN metastases and performed in those with >3 SLN metastases and/or gross extracapsular extension ^47^. Both cases with pure ILC and mixed ILC/NST were classified as ILC. After 3.5 years of follow-up, no significant differences were seen in recurrence and survival rates when comparing all ILC and all NST cases (independent of axillary management). In the present study, the ILC/NST mixed subtype was presented separately. Although no association with the risk of a high nodal burden was found, this finding has to be interpreted with caution due to the low prevalence of the ILC/NST mixed subtype.

Whether the findings of a higher axillary nodal burden in ILC after cALND in the present study have implications on clinical outcome needs to be further investigated. A multidisciplinary discussion regarding omission of cALND is encouraged for all patients with ILC.

## Data Availability

The dataset used during the current study are available from the corresponding author on reasonable request.

## Article information

### Author Contributions

All authors made substantial contributions to conception and design, acquisition of data, or analysis and interpretation of data; took part in drafting the article or revising it critically for important intellectual content. For details see attachment.

